# No evidence for causal effects of C-reactive protein (CRP) on chronic pain conditions: a Mendelian randomization study

**DOI:** 10.1101/2024.07.03.24309700

**Authors:** Pradeep Suri, Yakov A. Tsepilov, Elizaveta E. Elgaeva, Frances M. K. Williams, Maxim B. Freidin, Ian B. Stanaway

**Author notes:** **Corresponding author:** Pradeep Suri, MD, MS, Mailing Address: Rehabilitation Care Services, VA Puget Sound Health Care System, S-RCS-117, 1660 S. Columbian Way, Seattle, WA, 98108.

## Abstract

**Objective:** We conducted a Mendelian randomization (MR) study to examine causal associations of C-reactive protein (CRP) with (1) spinal pain; (2) extent of multisite chronic pain; and (3) chronic widespread musculoskeletal pain.

**Design:** Two-sample MR study.

**Setting/Subjects:** We used summary statistics from publicly available genome-wide association studies (GWAS) conducted in multiple cohorts and biobanks. Genetic instrumental variables were taken from an exposure GWAS of CRP (n=204,402). Outcome GWASs examined spinal pain (n=1,028,947), extent of multisite chronic pain defined as the number of locations with chronic pain (n=387,649), and chronic widespread pain (n=249,843).

**Methods:** We examined MR evidence for causal associations using inverse-variance weighted (IVW) analysis and sensitivity analyses using other methods. We calculated odds ratios (ORs), 95% confidence intervals (95% CIs), and p-values, using a Bonferroni correction (p<0.0166) to account for 3 primary comparisons.

**Results:** Greater serum CRP (mg/L) was not significantly causally associated with spinal pain (OR=1.04, 95% CI 1.00-1.08; p=0.07) in IVW analysis. Greater serum CRP also showed no significant causal association with extent of multisite chronic pain in IVW analysis (beta coefficient= 0.014, standard error=0.011; p=0.19). CRP also showed no significant causal association with chronic widespread pain in IVW analysis (OR=1.00, 95% CI 1.00-1.00; p=0.75). All secondary and sensitivity analyses also showed no significant associations.

**Conclusions:** This MR study found no causal association of CRP on spinal pain, the extent of chronic pain, or chronic widespread pain. Future studies examining mechanistic biomarkers for pain conditions should consider other candidates besides CRP.

## INTRODUCTION

Chronic pain affects an estimated 116 million adults in the United States and costs up to $635 billion each year.^1,2^ In response to this public health problem, the Federal Pain Research Strategy has promoted improvements in pain management.^3^ The discovery and validation of useful pain biomarkers is a critical step on the path towards finding better options for pain management. Biomarkers for pain mechanisms and chronic pain predisposition are two important categories of human pain biomarkers.^4^

Inflammatory biomarkers have been extensively studied in inflammatory musculoskeletal conditions associated with joint and spinal pain such as rheumatoid arthritis^5^ and spondyloarthritis,^6^ and some—such as C-reactive protein (CRP), an acute inflammatory protein expressed during inflammation and infection— are commonly used clinically. CRP is an immune regulator, rather than only a marker for inflammation or infection,^7^ so a potential causal role for CRP may exist in inflammatory musculoskeletal conditions. Given the potential and actual utility of CRP in inflammatory conditions, CRP has become one the most commonly studied biomarkers in non-inflammatory pain conditions.^8-10^ However, studies of associations between CRP and non-inflammatory pain conditions have had mixed results.^8-11^ Moreover, many studies of CRP as a biomarker for pain conditions have had limitations such as small sample sizes; not accounting for multiple statistical comparisons; limited adjustment for potential confounders; and cross-sectional designs in which only limited insight can be gained into temporality.^9,10 8,11^ The attention paid to CRP in prior studies of pain may be due to convenience, such as the widespread availability of CRP in clinical settings and the fact that CRP has existing clinical use for health conditions. Convenience is a less compelling reason to study a biomarker than a strong conceptual rationale regarding the precise underlying mechanisms of pain. Resources spent examining mechanistic biomarkers without a strong conceptual rationale may be more likely to show null findings and waste limited resources that might otherwise be directed towards other candidate biomarkers.^4,12^

With this in mind we used existing data to conduct a Mendelian randomization study to examine causal associations of CRP with three pain-related conditions: (1) spinal pain; (2) extent of multisite chronic pain ; and (3) chronic widespread pain.

## METHODS

A two-sample Mendelian randomization (MR) study was performed using publicly available genome-wide association study summary statistics (i.e., no individual-level data). MR uses single-nucleotide variants (or “SNVs”) as genetic “instrumental variables” to examine associations between an exposure and an outcome that are typically interpreted as causal associations. The strengths, limitations, and core assumptions of MR and instrumental variable analyses, including the relevance, independence, and exclusion restriction assumptions, have been well-described elsewhere. ^13^ Briefly, MR offers greater control for potential confounders than conventional observational studies due in part to the rigorous methods used to adjust for population stratification in genome-wide association studies (GWAS). While residual confounding can occur in GWAS for traits with strong psychosocial components,^14^ the potential for such confounding is generally lower when studying the causal effects of molecular phenotypes such as CRP.^15^ Additionally, MR studies allow a clear temporal sequence whereby genetic proxies for the exposure precede the outcome, minimizing potential bias due to reverse causation.^13^ Two-sample MR studies use two separate GWAS samples to determine causal exposure-outcome relationships: an exposure GWAS, which is used to identify the genetic instrumental variables (i.e., SNVs) that serve as genetic proxies for the exposure of interest; and an outcome GWAS. Importantly, the exposure and outcome GWAS used in two-sample MR studies should not overlap, as this can bias towards the conventional observational estimate.^13^ No ethics approvals were required because this work used publicly available summary statistics only. Analyses were completed between September 2022 and March 2023. We followed the STROBE-MR checklist to guide reporting.^16^

### Exposure GWAS of CRP and genetic instrumental variable selection

CRP was expected to yield strong instruments for MR studies because of its high additive SNV-heritability, estimated at 13%,^17^ and the low propensity of molecular phenotypes to produce biased or confounded MR associations.^15^ We selected SNV instrument variables that were associated with CRP at less than the conventional threshold of genome-wide significance (p<5×10^−8^) in a previously published meta-GWAS of CRP by Ligthart et al., conducted in 204,402 European participants.^18^ A polygenic risk score using these variants was significantly positively correlated with serum CRP in an independent sample.^19^ The meta-GWAS by Ligthart et al. comprised 88 cohorts from the Cohorts for Heart and Aging Research in Genomic Epidemiology (CHARGE) consortium. Analyses included only participants of European ancestry to limit between-sample confounding by ancestry differences.^13^ Cohorts in the meta-GWAS measured serum CRP in mg/L using standard assays and laboratory techniques,^18^ and CRP values were natural log-transformed for analysis. Individuals with autoimmune disease, those who were taking immune-modulating agents, and/or those with CRP outlier values more than 4 SD from the mean were excluded from analyses. The *F* statistic for 52 of the instruments identified by Ligthart et al. was 273, far exceeding the commonly used threshold of >10 used to identify strong instruments, supporting the relevance assumption.^13^ The *F* statistic for the single cis-variant among these instruments in the CRP gene (rs2794520) was 2,902.^13^ Analyses of association between genetic variants and CRP were conducted using an additive linear regression model adjusted for age, sex, and principal components. Variants were filtered based on imputation quality *R*^2^>0.4. Further details of the GWAS study of CRP are available elsewhere. ^18^

### Outcome GWAS of spinal pain, extent of multisite chronic pain, and chronic widespread pain

Extended details regarding genotyping, imputation, and analyses can be found in the parent study meta-GWAS of spinal pain, extent of multisite chronic pain, and chronic widespread pain.^20-22^ In brief, we used summary data from the largest available GWAS of the targeted pain phenotypes (spinal pain; extent of multisite chronic pain, and chronic widespread pain) that did not overlap with the exposure GWAS of CRP conducted in the CHARGE consortium. Genetic variants associated with spinal pain were identified from a meta-GWAS of spinal pain associated with healthcare use in 1,028,947 individuals of European ancestry (119,100 cases and 909,847 controls) from UK Biobank, the FinnGen project, Iceland, and Denmark.^20^ This study identified spinal pain cases using the M54 “dorsalgia” code group of the International Statistical Classification of Diseases, 10^th^ Revision (ICD-10). This phenotype reflects both back and neck pain requiring healthcare utilization, with or without associated radicular signs or symptoms. It includes both acute and chronic spinal pain but is expected to comprise mainly the latter due to the association of chronic back pain with health care seeking.^23-25^ Similar health record-defined spinal pain phenotypes are widely used in pain research^26-29^ and their validity is supported by replicable results using such phenotypes across countries and healthcare systems.^30^ Analyses of association between genetic variants and spinal pain in these biobanks generally adjusted for the same factors across biobanks, including age, sex, study-specific factors (e.g., batch), and principal components.

Variants associated with the extent of multisite chronic pain were identified from a GWAS conducted in 387,649 UK Biobank participants. ^21^ This study defined the extent of multisite chronic pain as a count of the number of locations in which pain had been present for ≥3 months, including the head, face, back, neck/shoulder, stomach/abdomen, hip, and knee. The number of chronic pain locations ranged from 0 to 7, with higher numbers reflecting greater extent of multisite chronic pain. Analyses of association between genetic variants and extent of multisite chronic pain adjusted for age, sex, array, and principal components.

Last, variants associated with chronic widespread pain were identified from a GWAS conducted in the UK Biobank including 6,914 chronic widespread pain cases and 242,929 controls. Chronic widespread pain cases were defined by having either (1) self-reported pain all over the body lasting ≥3 months; (2) self-reported simultaneous pain in the knee, shoulder, hip and back lasting ≥3 months ; and/or (3) self-reported or healthcare professional-diagnosed fibromyalgia.^22^ Chronic widespread pain controls were those who reported no pain in the last month; or reported pain all over the body in the previous month that did not last for 3 months; or reported ≥3 months of non-musculoskeletal pain only (headache and/or pain in the face or stomach/abdomen). Those reporting inflammatory arthritis, systemic lupus erythematosus, or myopathy were excluded. Analyses of association between genetic variants and chronic widespread pain adjusted for age, sex, array, and principal components.

### Statistical analysis

All analyses were performed in R v4.1.1. Exposure and outcome GWAS did not overlap, and the same set of covariates (age, sex, study-specific factors such as array, principal components) was used for adjustment in each exposure-outcome pair. To harmonize the exposure and outcome data alleles we used the “TwoSampleMR” R package *harmonise_data* function with the action=1 flag and inspected the instrumental variables for concordant allele identities. For each CRP-outcome trait pair we prepared a list of overlapping SNVs and performed clumping for independence of selected IVs within a 10000 kb window, using a genome-wide significance threshold p-value ≤ 5 × 10^−8^ for association, an r^2^[?]>[?]0.001 threshold for correlation, and excluding IVs with minor allele frequency (MAF) < 0.05. MR analyses were conducted using the “TwoSampleMR” R package *mr* function. The primary causal effect estimate for each risk factor and outcome pair was obtained through an inverse variance-weighted (IVW) meta-analysis of the ratios of the exposure effect size and outcome effect size for each variant. We calculated odds ratios (ORs), 95% confidence intervals (95% CIs), and p-values, using a Bonferroni correction (0.05/3=0.0166) to account for the 3 primary comparisons made for the associations of CRP with spinal pain, multisite chronic pain, and chronic widespread pain. Secondary analyses using other MR methods (MR Egger, weighted median, simple mode, and weighted mode) were also conducted to examine the robustness of findings. If MR Egger intercepts were statistically significant, suggesting possible directional horizontal pleiotropy, we tested for this using the *mr_pleiotropy_test* function. We calculated the magnitude of detectable odds ratios (ORs), assuming 80% power and p=0.0166, using methods we have described elsewhere.^31^ We also conducted sensitivity analyses using the SNV rs2794520 to examine whether associations would be different when restricted to a proximal cis-variant within 5kB of the CRP gene. Due to the strength of the CRP instruments used, and the known tendency for false positives produced by the MR estimators above,^32^ we planned to conduct further analyses using the CAUSE method if IVW and sensitivity analyses indicated causal associations, as we have in past work.^31,33^

## RESULTS

We identified 54 genetic instrumental variables for CRP. Very few instruments were associated with the pain condition outcomes at even nominal significance (p<0.05), and none attained genome-wide significance (Supplemental File 1). The main study findings are presented in Table 1. Greater units of serum CRP (mg/L) showed no significant causal associations with spinal pain (OR=1.04, 95% CI 1.00-1.08; p=0.07) in the IVW analysis. Secondary analyses using other MR methods (weighted median, simple mode, weight mode, and MR-Egger) also revealed no significant associations with spinal pain, with OR point estimates from different analytic methods that were both higher and lower than 1.0, without any consistent pattern of association (Table 1). There was no evidence of directional horizontal pleiotropy (MR-Egger intercept 0.002, standard error [SE]=0.002, p=0.28).

**Table 1.**
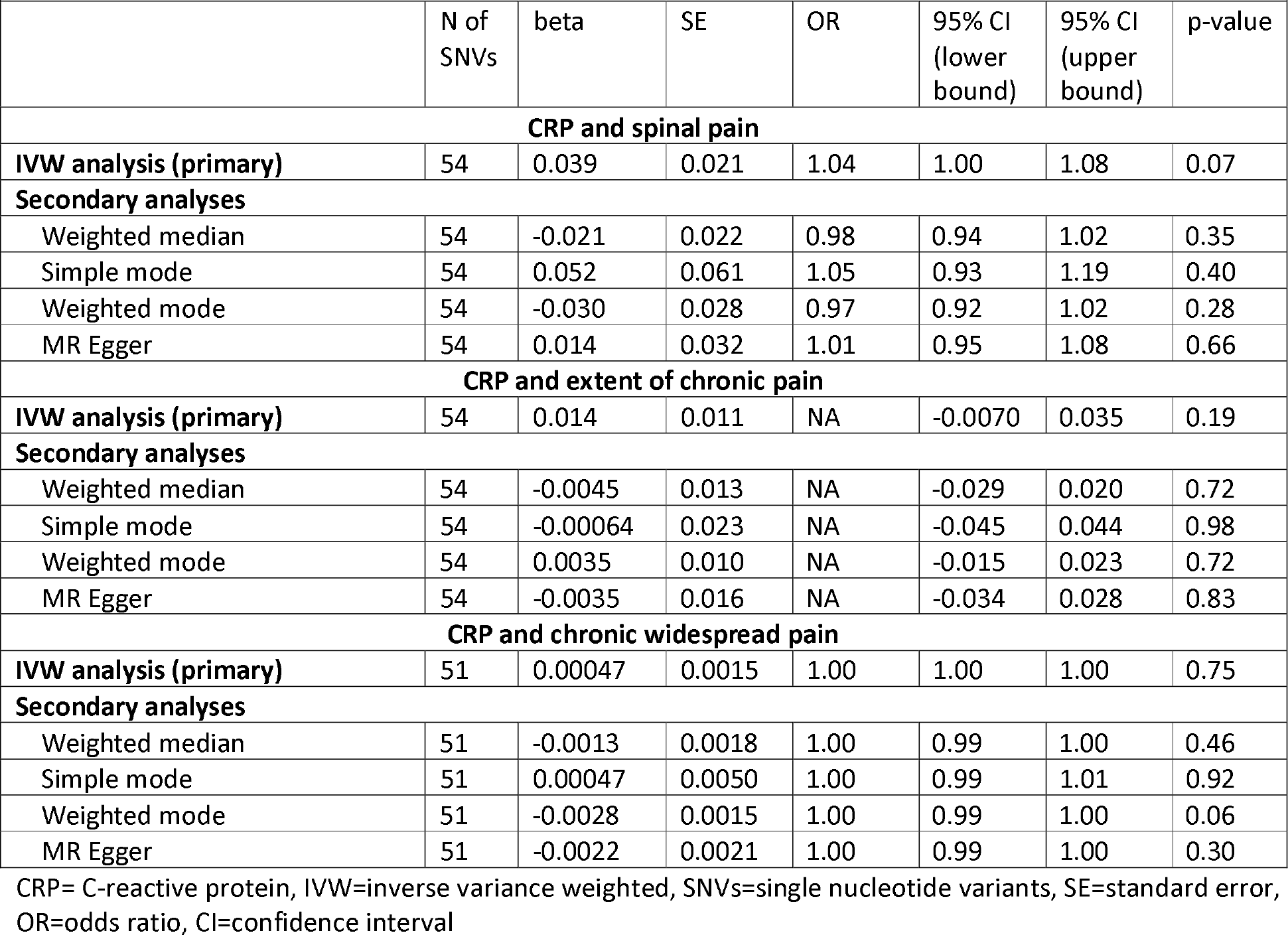
Causal associations of CRP with pain conditions.

| **Table 1. Causal associations of CRP with pain conditions** |  |  |  |  |  |  |  |
| --- | --- | --- | --- | --- | --- | --- | --- |
|  | N of SNVs | beta | SE | OR | 95% CI (lower bound) | 95% CI (upper bound) | p-value |
| **CRP and spinal pain** |  |  |  |  |  |  |  |
| **IVW analysis (primary)** | 54 | 0.039 | 0.021 | 1.04 | 1.00 | 1.08 | 0.07 |
| **Secondary analyses** |  |  |  |  |  |  |  |
| Weighted median | 54 | -0.021 | 0.022 | 0.98 | 0.94 | 1.02 | 0.35 |
| Simple mode | 54 | 0.052 | 0.061 | 1.05 | 0.93 | 1.19 | 0.40 |
| Weighted mode | 54 | -0.030 | 0.028 | 0.97 | 0.92 | 1.02 | 0.28 |
| MR Egger | 54 | 0.014 | 0.032 | 1.01 | 0.95 | 1.08 | 0.66 |
| **CRP and extent of chronic pain** |  |  |  |  |  |  |  |
| **IVW analysis (primary)** | 54 | 0.014 | 0.011 | NA | -0.0070 | 0.035 | 0.19 |
| **Secondary analyses** |  |  |  |  |  |  |  |
| Weighted median | 54 | -0.0045 | 0.013 | NA | -0.029 | 0.020 | 0.72 |
| Simple mode | 54 | -0.00064 | 0.023 | NA | -0.045 | 0.044 | 0.98 |
| Weighted mode | 54 | 0.0035 | 0.010 | NA | -0.015 | 0.023 | 0.72 |
| MR Egger | 54 | -0.0035 | 0.016 | NA | -0.034 | 0.028 | 0.83 |
| **CRP and chronic widespread pain** |  |  |  |  |  |  |  |
| **IVW analysis (primary)** | 51 | 0.00047 | 0.0015 | 1.00 | 1.00 | 1.00 | 0.75 |
| **Secondary analyses** |  |  |  |  |  |  |  |
| Weighted median | 51 | -0.0013 | 0.0018 | 1.00 | 0.99 | 1.00 | 0.46 |
| Simple mode | 51 | 0.00047 | 0.0050 | 1.00 | 0.99 | 1.01 | 0.92 |
| Weighted mode | 51 | -0.0028 | 0.0015 | 1.00 | 0.99 | 1.00 | 0.06 |
| MR Egger | 51 | -0.0022 | 0.0021 | 1.00 | 0.99 | 1.00 | 0.30 |

CRP= C-reactive protein, IVW=inverse variance weighted, SNVs=single nucleotide variants, SE=standard error, OR=odds ratio, CI=confidence interval

Greater units of serum CRP in mg/L showed no significant causal associations with extent of multisite chronic pain as measured by increasing number of chronic pain locations in the IVW analyses (beta coefficient= 0.014, standard error=0.011; p=0.19). Secondary analyses using other MR methods also showed no significant associations, with OR point estimates both higher and lower than 0 and no consistent pattern of association (Table 1). There was no evidence of directional horizontal pleiotropy (MR-Egger intercept 0.001, standard error [SE]=0.001, p=0.14).

Greater units of serum CRP in mg/L showed no significant causal associations with chronic widespread pain in the IVW analysis (OR=1.00, 95% CI 1.00-1.00; p=0.75). Secondary analyses using other MR methods again showed no significant associations, with OR point estimates both higher and lower than 0 and no consistent pattern of association (Table 1). There was no significant evidence of directional horizontal pleiotropy (MR-Egger intercept 0.0002, standard error [SE]=0.0001, p=0.09).

Sensitivity analyses using the cis-SNV rs2794520 that was localized to the CRP gene also showed no significant causal associations with spinal pain (OR=0.97, 95% CI 0.92-1.03; p=0.31), number of chronic pain locations (beta coefficient= -0.019, standard error=0.014; p=0.19) or chronic widespread pain (OR=1.00, 95% CI 0.99-1.01; p=0.10).

*Post hoc* power calculations indicated 80% power to detect an odds ratio of 1.03 in the MR analyses of spinal pain, 1.02 in the analyses of extent of chronic pain, and 1.11 in the analyses of chronic widespread pain. Given the consistency of null associations across all analyses conducted, no additional analyses were conducted using the CAUSE method.

## DISCUSSION

This two-sample Mendelian randomization study analyzed data on CRP from 204,402 participants and data from between 249,843 to 1,028,947 participants for the outcomes of spinal pain, extent of chronic pain, or chronic widespread pain. Despite the large samples involved, no significant evidence was found for a causal association of CRP with these pain conditions.

This study’s finding of no causal association of CRP with spinal pain is convincing and likely robust in light of the very strong instruments, with F-statistics of 273 and 2,902 for all SNV instruments and the cis-variant rs2794520, respectively, that were yielded by the exposure GWAS of CRP.^13^ This instrument strength and the SNV-heritability it mirrors far exceeds that of instruments which our group has used previously for other risk factors and been able to detect significant causal MR associations.^31,33,34^ Our null finding for the CRP-spinal pain association is consistent with the mixed prior results from small observational studies of the related phenotype of low back pain, including two studies finding no significant associations with chronic low back pain,^35,36^ one study finding significantly greater CRP levels in acute back pain cases versus controls,^37^ and two studies finding higher levels of CRP in whiplash-associated disorder cases vs. controls.^38,39^ Importantly, the small size of most prior observational studies, typically with case-control designs and with fewer than 50 individuals per arm,^35,38,39^ provided limited ability to control for potential confounders. The importance of controlling for potential confounders is illustrated by a recent, large-scale cross-sectional analysis also conducted in UK Biobank, which found a moderate-magnitude CRP-low back pain association in unadjusted analyses (OR=1.37, 95% CI 1.36-1.39) of females which was markedly attenuated after adjustment for a relatively small number of covariates (OR=1.08, 95% CI 1.06-1.10) given the large sample size available and the numerous potential confounders available for covariate adjustment in UK Biobank.^40^ This study found the same trend of markedly attenuated associations of CRP with pain after covariate adjustment across all pain locations and types, and in both sexes, strongly suggesting that further accounting for potential confounders would have further reduced the CRP-pain relationship to a null association.^40^ Future large-scale non-MR studies attempting to remove residual confounding affecting the CRP-pain relationship should select covariates informed by causal diagrams, avoiding adjustment for potential colliders;^41^ should include multiple inflammatory and infectious conditions as separate adjustment variables, rather than general comorbidity scores; and should use longitudinal designs, which can have lower potential for inducing collider bias via adjustment for *consequences* of CRP and pain if the choice of adjustment variables is appropriate and selection not induced when defining the cohort.^41^ Taken together, the prior literature on the CRP-low back pain association reminds us that associations from cross-sectional and case-control studies are low on the hierarchy of scientific evidence, and the majority of associations found in such studies are not likely to be causal. The current study, allowing much greater control for confounding through the MR design and focusing on causal associations, found a robustly null association between CRP and low back pain.

While our study finding no causal association of CRP was the first MR examining this topic for the outcome of spinal pain, its results fit with those from the 3 previous MR studies of back pain. Importantly, these were two-sample MR studies using outcome GWASs with sample sizes <500,000 participants, less than half the size of the outcome GWAS of spinal pain (n=1,028,947) used in the current study. One MR of CRP and back pain found a statistically significant association in IVW analyses,^42^ but the authors noted that this result might be explained by horizontal pleiotropy, or genetic confounding. Another two-sample MR found both significant and non-significant associations between CRP and back pain (and a mixed neck pain/shoulder pain phenotype), but the authors pointed out that the statistically significant associations may be due to sample overlap, which was present and could have biased towards the confounded observational estimate.^43^ The most recent GWAS of CRP and chronic back pain found no causal association.^44^ It has been argued that a major role for MR is for establishing *negative* results, as while biases such as confounding can in theory bias effect estimates towards the null, in actual practice confounding is more likely to be a problem when effect estimates *do* indicate an effect^45^ (rather than when they indicate *no* causal effect such as in the current study). Overall, given the very strong CRP instruments we used, large sample sizes, and the known tendency of the MR estimators we used to produce false *positive* findings^32^ (as opposed to false *negatives*), the current findings taken in the context of prior observational and MR studies do not support the existence of causal effects of CRP on spinal pain.^44^ Given that our spinal pain phenotype includes both acute and chronic cases of spinal pain, and acute pain is a necessary though not sufficient precursor to chronic pain, a causal effect of CRP on either acute or chronic spinal pain seems unlikely. This likely rules out CRP as a mechanistic biomarker but does not preclude CRP having utility as a prognostic biomarker.

To our knowledge, no prior MR studies have examined CRP as a potential cause of the extent of chronic pain or chronic widespread pain. A systematic review of observational studies examining CRP and fibromyalgia found five studies reporting higher levels of CRP among patients with fibromyalgia and 3 studies reporting no differences, with a statistically significant difference in CRP between cases vs. controls in meta-analysis.^8^ However the authors of this work cautioned that their findings “*cannot support the notion that these blood biomarkers are specific biomarkers to or diagnostic of* [fibromyalgia]” due the fact that CRP (and the other biomarkers studied) are also significantly different in cases (vs. controls) for many other pain and non-pain conditions, which themselves are associated with fibromyalgia.^8^ This reflects the basic issues of confounding and lack of specificity that may be problematic in observational studies of CRP’s association with pain. The same problem may affect other inflammatory biomarkers. Conventional observational studies, even if very large, may be fundamentally challenged to pinpoint causal associations of CRP on pain conditions unless they are longitudinal, as cross-sectional studies are not able to control for many potential confounders without potentially inducing collider bias.

CRP is among the most commonly studied biomarkers in chronic pain, and this may reflect its widespread availability in clinical care. However, convenience is not a compelling reason to study a biomarker. A systematic review of observational and MR studies across diverse phenotypes concluded that despite the vast amount of research dedicated to CRP, substantial support does not exist for CRP as a causal factor for *any* phenotype.^46^ With regards to the search for pain biomarkers specifically, agnostic approaches to identifying novel biomarkers may be a more fruitful way to identify mechanistic biomarkers for pain conditions. Such approaches will be challenging using the traditional model of analyzing case-control clinical samples, as multiple statistical testing corrections and scans across hundreds of candidate markers will require many thousands of participants per arm. A more fruitful strategy that is likely less prone to false positives, and one readily achievable using existing technology and publicly available resources, is to use large-scale genomic biobanks for mechanistic biomarker discovery in sample sizes of hundreds of thousands of participants, and to follow-up the top associations in conventional case-control or cohort studies.

A limitation of the current study is that it is only informative as to causal or mechanistic biomarkers, and it cannot inform as to whether CRP can provide prognostic information that is not causal. CRP may very well have value as a purely prognostic biomarker for pain conditions. Of note, while there was abundant power to detect small ORs for the analyses of spinal pain and extent of chronic pain, power was somewhat limited in the analysis of chronic widespread pain, with our *post hoc* calculations indicating that causal effects of CRP smaller than OR=1.11 may not have been detected. Another potential limitation of this study is that we did not conduct MR analyses using newer MR methods such as latent causal variable approaches, such as Causal Analysis Using Summary Effect estimates (CAUSE). Such approaches generally have less precision than the MR methods used in the current analysis, however, so they would be unlikely to have shown statistically significant results. Moreover, CAUSE was developed to mitigate the general tendency for the MR methods used in the current study to produce too many *false positive* results,^32^ which was quite the opposite of what we found in the current study with a remarkably consistent pattern of null associations. Nevertheless, future MR studies of CRP-pain relationships using larger samples may consider other analytic approaches that might be more robust to genetic confounding. Another limitation is that the current analyses included European participants only, which was done to decrease heterogeneity that might bias MR effect estimates; future studies should examine diverse samples.

In summary, this MR study found no significant causal association of CRP on spinal pain, the extent of chronic pain, or chronic widespread pain. Future studies examining mechanistic biomarkers for pain conditions should also consider other candidates besides CRP.

## Supporting information

STROBE MR checklist

## Data Availability

All data used in this manuscript are already publicly available. Websites where the summary statistics used can be found are provided in the "data availability links" section.

https://www.decode.com/summarydata/

https://www.cell.com/ajhg/fulltext/S0002-9297(18)30320-3

https://doi.org/10.5281/zenodo.4459546

http://dx.doi.org/10.5525/gla.researchdata.822.

## ACKNOWLEDGEMENTS

Drs. Suri and Stanaway are supported by VA I01RX0004291. Dr. Suri is an employee of the VA Puget Sound Health Care System and Director of the Resource Core of the University of Washington Clinical Learning, Evidence and Research (CLEAR) Center, which is funded by NIAMS/NIH P30AR072572. Ms. Elgaeva is supported by the Ministry of Education and Science of the RF via the Institute of Cytology and Genetics (project 0259-2021-0009 / AAAA-A17-117092070032-4). The contents of this work do not represent the views of the US Department of Veterans Affairs, the National Institutes of Health, or the US Government. The study was conducted using publicly available summary data from the studies mentioned above, and we are grateful for the study participants for making such research possible. protocol. FW is supported by Versus Arthritis grant 22467. The authors have no conflicts of interest.

## DATA AVAILABILITY

This study was conducted using publicly available summary data.

## Notes

**Funding Source:** Drs. Suri and Stanaway are supported by VA I01RX0004291. Dr. Suri is an employee of the VA Puget Sound Health Care System and Director of the Resource Core of the University of Washington Clinical Learning, Evidence and Research (CLEAR) Center, which is funded by NIAMS/NIH P30AR072572. Dr. Tsepilov is supported by the Wellcome Sanger Institute and the Russian Foundation for Basic Research (project 19-015-00151). Ms. Elgaeva is supported by the Ministry of Education and Science of the Russian Foundation via the Institute of Cytology and Genetics (project 0259-2021-0009 / AAAA-A17-117092070032-4). The contents of this work are solely the responsibility of the authors and do not represent the views of the US Department of Veterans Affairs, the National Institutes of Health, or the US Government. The study was conducted using publicly available summary data from the studies mentioned above, and we are grateful for the study participants for making such research possible.

**Potential Conflicts of Interest:** None of the authors has potential conflicts of interest to report.

### Competing Interest Statement

The authors have declared no competing interest.

### Author Declarations

Summary statistics for the four genome-wide association studies used in this analyses were publicly available for download prior to analyses.

