## Supplementary material for "No evidence for causal effects of C-reactive protein (CRP) on chronic pain conditions: a Mendelian randomization study": STROBE MR checklist

**STROBE-MR checklist of recommended items to address in reports of Mendelian randomization studies**^1^ ^2^

| **Item No.** | **Section** | **Checklist item** | **Page No.** | **Relevant text from manuscript** |
| --- | --- | --- | --- | --- |
| 1 | **TITLE and ABSTRACT** | Indicate Mendelian randomization (MR) as the study’s design in the title and/or the abstract if that is a main purpose of the study | Title | “No evidence for causal effects of C-reactive protein (CRP) on chronic pain conditions: a Mendelian randomization study” |
|  | **INTRODUCTION** |  |  |  |
| 2 | **Background** | Explain the scientific background and rationale for the reported study. What is the exposure? Is a potential causal relationship between exposure and outcome plausible? Justify why MR is a helpful method to address the study question | 1 | Inflammatory biomarkers have been extensively studied in inflammatory musculoskeletal conditions associated with joint and spinal pain such as rheumatoid arthritis^5^ and spondyloarthritis,^6^ and some—such as C-reactive protein (CRP), an acute inflammatory protein expressed during inflammation and infection— are commonly used clinically. CRP is an immune regulator, rather than only a marker for inflammation or infection,^7^ so a potential causal role for CRP may exist in inflammatory musculoskeletal conditions. Given the potential and actual utility of CRP in inflammatory conditions, CRP has also become among the most commonly studied biomarkers in non-inflammatory pain conditions.^8-10^ |
| 3 | **Objectives** | State specific objectives clearly, including pre-specified causal hypotheses (if any). State that MR is a method that, under specific assumptions, intends to estimate causal effects | 1 | “… we used existing data to conduct a Mendelian randomization study to examine causal associations of CRP with three pain-related conditions: (1) spinal pain; (2) extent of multisite chronic pain ; and (3) chronic widespread pain. ” |
|  | **METHODS** |  |  |  |
| 4 | **Study design and data sources** | Present key elements of the study design early in the article. Consider including a table listing sources of data for all phases of the study. For each data source contributing to the analysis, describe the following: |  |  |
|  | a) | Setting: Describe the study design and the underlying population, if possible. Describe the setting, locations, and relevant dates, including periods of recruitment, exposure, follow-up, and data collection, when available. | 2-3 | Sections “*Exposure GWAS of CRP and genetic instrumental variable selection*”, “*Outcome GWAS of spinal pain, extent of multisite chronic pain, and chronic widespread pain*”, and the provided citations in these sections |
|  | b) | Participants: Give the eligibility criteria, and the sources and methods of selection of participants. Report the sample size, and whether any power or sample size calculations were carried out prior to the main analysis | 2-3 | Sections “*Exposure GWAS of CRP and genetic instrumental variable selection*”, “*Outcome GWAS of spinal pain, extent of multisite chronic pain, and chronic widespread pain*”, and the provided citations in these sections |
|  | c) | Describe measurement, quality control and selection of genetic variants | 2-3 | Sections “*Exposure GWAS of CRP and genetic instrumental variable selection*”, “*Outcome GWAS of spinal pain, extent of multisite chronic pain, and chronic widespread pain*”, and the provided citations in these sections |
|  | d) | For each exposure, outcome, and other relevant variables, describe methods of assessment and diagnostic criteria for diseases | 2-3 | Sections “*Exposure GWAS of CRP and genetic instrumental variable selection*”, “*Outcome GWAS of spinal pain, extent of multisite chronic pain, and chronic widespread pain*”, and the provided citations in these sections |
|  | e) | Provide details of ethics committee approval and participant informed consent, if relevant | 2 | “No ethics approvals were required because this work used publicly available summary statistics only. ” |
| 5 | **Assumptions** | Explicitly state the three core IV assumptions for the main analysis (relevance, independence and exclusion restriction) as well assumptions for any additional or sensitivity analysis | 1 | The strengths, limitations, and core assumptions of MR and instrumental variable analyses, including the relevance, independence, and exclusion restriction assumptions, have been well-described elsewhere. ^13^ |
| 6 | **Statistical methods: main analysis** | Describe statistical methods and statistics used |  |  |
|  | a) | Describe how quantitative variables were handled in the analyses (i.e., scale, units, model) | 2 | Cohorts in the meta-GWAS measured serum CRP in mg/L using standard assays and laboratory techniques,^18^ and CRP values were natural log-transformed for analysis. Individuals with autoimmune disease, those who were taking immune-modulating agents, and/or those with CRP outlier values more than 4 SD from the mean were excluded from analyses. |
|  | b) | Describe how genetic variants were handled in the analyses and, if applicable, how their weights were selected | 3 | Sections “*Exposure GWAS of CRP and genetic instrumental variable selection*”, “*Outcome GWAS of spinal pain, extent of multisite chronic pain, and chronic widespread pain*” and “*Statistical analysis*” |
|  | c) | Describe the MR estimator (e.g. two-stage least squares, Wald ratio) and related statistics. Detail the included covariates and, in case of two-sample MR, whether the same covariate set was used for adjustment in the two samples | 3 | Section “*Statistical analysis*”, “The primary causal effect estimate for each risk factor and outcome pair was obtained through an inverse variance-weighted (IVW) meta-analysis of the ratios of the exposure effect size and outcome effect size for each variant. We calculated odds ratios (ORs), 95% confidence intervals (95% CIs), and p-values, using a Bonferroni correction (0.05/3=0.0166) to account for the 3 primary comparisons made for the associations of CRP with spinal pain, multisite chronic pain, and chronic widespread pain. Secondary analyses using other MR methods (MR Egger, weighted median, simple mode, and weighted mode) were also conducted to examine the robustness of findings. If MR Egger intercepts were statistically significant, suggesting possible directional horizontal pleiotropy, we tested for this using the *mr_pleiotropy_test* function. We calculated the magnitude of detectable odds ratios (ORs), assuming 80% power and p=0.0166, using methods we have described elsewhere.[34] We also conducted sensitivity analyses using the SNV rs2794520 to examine whether associations would be different when restricted to cis-variants in the CRP gene.”  “Exposure and outcome GWAS did not overlap, and the same set of covariates (age, sex, study-specific factors such as array, principal components) was used for adjustment in each exposure-outcome pair. ” |
|  | d) | Explain how missing data were addressed | 2 | Sections “*Exposure GWAS of CRP and genetic instrumental variable selection*” (“Variants were filtered based on imputation quality *R*^2^>0.4. Further details of the GWAS study of CRP are available elsewhere. [25] ”), “*Outcome GWAS of spinal pain, extent of multisite chronic pain, and chronic widespread pain*” (“*Extended details regarding genotyping, imputation, and analyses can be found in the parent study meta-GWAS of spinal pain, extent of multisite chronic pain, and chronic widespread pain.[2; 20; 34]*”) |
|  | e) | If applicable, indicate how multiple testing was addressed |  | “We calculated odds ratios (ORs), 95% confidence intervals (95% CIs), and p-values, using a Bonferroni correction (0.05/3=0.0166) to account for the 3 primary comparisons made for the associations of CRP with spinal pain, multisite chronic pain, and chronic widespread pain. ” |
| 7 | **Assessment of assumptions** | Describe any methods or prior knowledge used to assess the assumptions or justify their validity | 2, 4, 5 | “The *F* statistic for the 52 instruments identified by Ligthart et al. was 273, far exceeding the commonly used threshold of >10 used to identify strong instruments, supporting the relevance assumption.^13^ The *F* statistic for the single cis-variant among these instruments in the CRP gene (rs2794520) was 2,902.^13^” “There was no significant evidence of directional horizontal pleiotropy using MR Egger(data not shown).” ”Due to the strength of the CRP instruments used, and the known tendency for false positives produced by the MR estimators above,^31^ we planned to conduct further analyses using the CAUSE method if IVW and sensitivity analyses indicated causal associations, as we have in past work.^30,32^ “Given the consistency of null associations across all analyses conducted, no additional analyses were conducted using the CAUSE method.” |
| 8 | **Sensitivity analyses and additional analyses** | Describe any sensitivity analyses or additional analyses performed (e.g. comparison of effect estimates from different approaches, independent replication, bias analytic techniques, validation of instruments, simulations) | 4 | “Secondary analyses using other MR methods (MR Egger, weighted median, simple mode, and weighted mode) were also conducted to examine the robustness of findings. If MR Egger intercepts were statistically significant, suggesting possible directional horizontal pleiotropy, we tested for this using the *mr_pleiotropy_test* function. We calculated the magnitude of detectable odds ratios (ORs), assuming 80% power and p=0.0166, using methods we have described elsewhere.^30^ We also conducted sensitivity analyses using the SNV rs2794520 to examine whether associations would be different when restricted to a proximal cis-variant within 5kB of the CRP gene. Due to the strength of the CRP instruments used, and the known tendency for false positives produced by the MR estimators above,^31^ we planned to conduct further analyses using the CAUSE method if IVW and sensitivity analyses indicated causal associations, as we have in past work.^30,32^” “Given the consistency of null associations across all analyses conducted, no additional analyses were conducted using the CAUSE method.” |
| 9 | **Software and pre-registration** |  |  |  |
|  | a) | Name statistical software and package(s), including version and settings used | 5 | “All analyses were performed in R v4.1.1.” |
|  | b) | State whether the study protocol and details were pre-registered (as well as when and where) | NA | We did not pre-register the protocol |
|  | **RESULTS** |  |  |  |
| 10 | **Descriptive data** |  |  |  |
|  | a) | Report the numbers of individuals at each stage of included studies and reasons for exclusion. Consider use of a flow diagram | 2-3 | Sections “*Exposure GWAS of CRP and genetic instrumental variable selection*”, “*Outcome GWAS of spinal pain, extent of multisite chronic pain, and chronic widespread pain*”, and the provided citations in these sections |
|  | b) | Report summary statistics for phenotypic exposure(s), outcome(s), and other relevant variables (e.g. means, SDs, proportions) | 2-3 | Summary statistics for the samples are provided in the citations included in the subsections “*Exposure GWAS of CRP and genetic instrumental variable selection*”, “*Outcome GWAS of spinal pain, extent of multisite chronic pain, and chronic widespread pain*” |
|  | c) | If the data sources include meta-analyses of previous studies, provide the assessments of heterogeneity across these studies | 2 | “Extended details regarding genotyping, imputation, and analyses can be found in the parent study meta-GWAS of spinal pain, extent of multisite chronic pain, and chronic widespread pain.^19-21^” |
|  | d) | For two-sample MR:  i.  Provide justification of the similarity of the genetic variant-exposure associations between the exposure and outcome samples  ii.  Provide information on the number of individuals who overlap between the exposure and outcome studies | 2. 3 | “In brief, we used summary data from the largest available GWAS of the targeted pain phenotypes (spinal pain; extent of multisite chronic pain, and chronic widespread pain) that did not overlap with the exposure GWAS of CRP conducted in the CHARGE consortium.” “Exposure and outcome GWAS did not overlap, ” |
| 11 | **Main results** |  |  |  |
|  | a) | Report the associations between genetic variant and exposure, and between genetic variant and outcome, preferably on an interpretable scale | 3, 4 | “A polygenic risk score using these variants has been significantly positively correlated with serum CRP in an independent sample.^19^ ”  “Very few instruments were associated with the pain condition outcomes at even nominal significance (p<0.05), and none attained genome-wide significance (Supplemental File 1). ” |
|  | b) | Report MR estimates of the relationship between exposure and outcome, and the measures of uncertainty from the MR analysis, on an interpretable scale, such as odds ratio or relative risk per SD difference |  | “The main study findings are presented in Table 1. Greater units of serum CRP (mg/L) showed no significant causal associations with spinal pain (OR=1.04, 95% CI 1.00-1.08; p=0.07) in the IVW analysis. Secondary analyses using other MR methods also revealed no significant associations with spinal pain, with OR point estimates from different analytic methods that were both higher and lower than 1.0, and no consistent pattern of association. Greater units of serum CRP in mg/L also showed no significant causal associations with extent of multisite chronic pain as measured by increasing number of chronic pain locations in the IVW analyses (beta coefficient= 0.014, standard error=0.011; p=0.19). Secondary analyses using other MR methods showed no significant associations, with OR point estimates both higher and lower than 0 and no consistent pattern of association (Table 1). Greater units of serum CRP in mg/L also showed no significant causal associations with chronic widespread pain in the IVW analysis (OR=1.00, 95% CI 1.00-1.00; p=0.75). ” |
|  | c) | If relevant, consider translating estimates of relative risk into absolute risk for a meaningful time period |  | NA, this was not relevant |
|  | d) | Consider plots to visualize results (e.g. forest plot, scatterplot of associations between genetic variants and outcome versus between genetic variants and exposure) |  | This was not relevant given the robustly null association |
| 12 | **Assessment of assumptions** |  |  |  |
|  | a) | Report the assessment of the validity of the assumptions | 2, 4, 5 | “The *F* statistic for the 52 instruments identified by Ligthart et al. was 273, far exceeding the commonly used threshold of >10 used to identify strong instruments, supporting the relevance assumption.^13^ The *F* statistic for the single cis-variant among these instruments in the CRP gene (rs2794520) was 2,902.^13^” “  “Secondary analyses using other MR methods (weighted median, simple mode, weight mode, and MR-Egger) also revealed no significant associations with spinal pain, with OR point estimates from different analytic methods that were both higher and lower than 1.0, without any consistent pattern of association. There was no evidence of directional horizontal pleiotropy (MR-Egger intercept 0.002, standard error [SE]=0.002, p=0.28).”  “Secondary analyses using other MR methods also showed no significant associations, with OR point estimates both higher and lower than 0 and no consistent pattern of association (Table 1). There was no evidence of directional horizontal pleiotropy (MR-Egger intercept 0.001, standard error [SE]=0.001, p=0.14).”  “Secondary analyses using other MR methods again showed no significant associations, with OR point estimates both higher and lower than 0 and no consistent pattern of association (Table 1). There was no significant evidence of directional horizontal pleiotropy (MR-Egger intercept 0.0002, standard error [SE]=0.0001, p=0.09).”  ”Due to the strength of the CRP instruments used, and the known tendency for false positives produced by the MR estimators above,^31^ we planned to conduct further analyses using the CAUSE method if IVW and sensitivity analyses indicated causal associations, as we have in past work.^30,32^ “Given the consistency of null associations across all analyses conducted, no additional analyses were conducted using the CAUSE method.” |
|  | b) | Report any additional statistics (e.g., assessments of heterogeneity across genetic variants, such as *I^2^*, Q statistic or E-value) | NA | This was not indicated given the robustly null findings above in row 12a |
| 13 | **Sensitivity analyses and additional analyses** |  |  |  |
|  | a) | Report any sensitivity analyses to assess the robustness of the main results to violations of the assumptions | NA | This was not indicated given the robustly null findings above in row 12a |
|  | b) | Report results from other sensitivity analyses or additional analyses | NA | This was not indicated given the robustly null findings above in row 12a |
|  | c) | Report any assessment of direction of causal relationship (e.g., bidirectional MR) | NA | This was not indicated given the unidirectional research question and the robustly null findings |
|  | d) | When relevant, report and compare with estimates from non-MR analyses | NA | This was not indicated given the study findings |
|  | e) | Consider additional plots to visualize results (e.g., leave-one-out analyses) | NA | This was not indicated given the study findings |
|  | **DISCUSSION** |  |  |  |
| 14 | **Key results** | Summarize key results with reference to study objectives | 5-6, paragraphs 1-4 of Discussion section | “This two-sample Mendelian randomization study analyzed data on CRP from 204,402 participants and data from between 249,843 to 1,028,947 participants for the outcomes of spinal pain, extent of chronic pain, or chronic widespread pain. Despite the large samples involved, no significant evidence was found for a causal association of CRP with these pain conditions. ” |
| 15 | **Limitations** | Discuss limitations of the study, taking into account the validity of the IV assumptions, other sources of potential bias, and imprecision. Discuss both direction and magnitude of any potential bias and any efforts to address them | p7, paragraph 6 of discussion section | “A limitation of the current study is that it is only informative as to causal or mechanistic biomarkers, and it cannot inform as to whether CRP can provide prognostic information that is not causal. CRP may very well have value as a purely prognostic biomarker for pain conditions. Of note, while there was abundant power to detect small ORs for the analyses of spinal pain and extent of chronic pain, power was somewhat limited in the analysis of chronic widespread pain, with our *post hoc* calculations indicating that causal effects of CRP smaller than OR=1.11 may not have been detected. Another potential limitation of this study is that we did not conduct MR analyses using newer MR methods such as latent causal variable approaches, such as Causal Analysis Using Summary Effect estimates (CAUSE). Such approaches generally have less precision than the MR methods used in the current analysis, however, so they would be unlikely to have shown statistically significant results. Moreover, CAUSE was developed to mitigate the general tendency for the MR methods used in the current study to produce too many *false positive* results,^32^ which was quite the opposite of what we found in the current study with a remarkably consistent pattern of null associations. Nevertheless, future MR studies of CRP-pain relationships using larger samples may consider other analytic approaches that might be more robust to genetic confounding. Another limitation is that the current analyses included European participants only, which was done to decrease heterogeneity that might bias MR effect estimates; future studies should examine diverse samples .” |
| 16 | **Interpretation** |  |  |  |
|  | a) | Meaning: Give a cautious overall interpretation of results in the context of their limitations and in comparison with other studies | 5-6, paragraphs 1-4 of Discussion section |  |
|  | b) | Mechanism: Discuss underlying biological mechanisms that could drive a potential causal relationship between the investigated exposure and the outcome, and whether the gene-environment equivalence assumption is reasonable. Use causal language carefully, clarifying that IV estimates may provide causal effects only under certain assumptions | NA | No causal mechanism was suggested |
|  | c) | Clinical relevance: Discuss whether the results have clinical or public policy relevance, and to what extent they inform effect sizes of possible interventions | P7 | “CRP is among the most commonly studied biomarkers in chronic pain, and this may reflect its widespread availability in clinical care. However, convenience is not a compelling reason to study a biomarker. A systematic review of observational and MR studies across diverse phenotypes concluded that despite the vast amount of research dedicated to CRP, substantial support does not exist for CRP as a causal factor for *any* phenotype.^45^ ” |
| 17 | **Generalizability** | Discuss the generalizability of the study results (a) to other populations, (b) across other exposure periods/timings, and (c) across other levels of exposure | P7 | “Another limitation is that the current analyses included European participants only, which was done to decrease heterogeneity that might bias MR effect estimates; future studies should examine diverse samples .” |
|  | **OTHER INFORMATION** |  |  |  |
| 18 | **Funding** | Describe sources of funding and the role of funders in the present study and, if applicable, sources of funding for the databases and original study or studies on which the present study is based | P7-8 | “Drs. Suri and Stanaway are supported by VA I01RX0004291. Dr. Suri is an employee of the VA Puget Sound Health Care System and Director of the Resource Core of the University of Washington Clinical Learning, Evidence and Research (CLEAR) Center, which is funded by NIAMS/NIH P30AR072572. Ms. Elgaeva is supported by the Ministry of Education and Science of the RF via the Institute of Cytology and Genetics (project 0259-2021-0009 / АААА-А17-117092070032-4). The contents of this work do not represent the views of the US Department of Veterans Affairs, the National Institutes of Health, or the US Government. The study was conducted using publicly available summary data from the studies mentioned above, and we are grateful for the study participants for making such research possible. protocol. FW is supported by Versus Arthritis grant 22467. The authors have no conflicts of interest.” |
| 19 | **Data and data sharing** | Provide the data used to perform all analyses or report where and how the data can be accessed, and reference these sources in the article. Provide the statistical code needed to reproduce the results in the article, or report whether the code is publicly accessible and if so, where | NA | This work only used publicly available data |
| 20 | **Conflicts of Interest** | All authors should declare all potential conflicts of interest | Provided with submission | None of the authors has potential conflicts of interest to report. |

This checklist is copyrighted by the Equator Network under the Creative Commons Attribution 3.0 Unported (CC BY 3.0) license.

1.	Skrivankova VW, Richmond RC, Woolf BAR, Yarmolinsky J, Davies NM, Swanson SA, et al. Strengthening the Reporting of Observational Studies in Epidemiology using Mendelian Randomization (STROBE-MR) Statement. JAMA. 2021;under review.

2.	Skrivankova VW, Richmond RC, Woolf BAR, Davies NM, Swanson SA, VanderWeele TJ, et al. Strengthening the Reporting of Observational Studies in Epidemiology using Mendelian Randomisation (STROBE-MR): Explanation and Elaboration. BMJ. 2021;375:n2233.
